# Is a COVID19 Quarantine Justified in Chile or USA Right Now?

**DOI:** 10.1101/2020.03.23.20042002

**Authors:** R. I. Gonzalez, F. Munoz, P. S. Moya, M. Kiwi

**Affiliations:** Centro de Nanotecnología Aplicada, Universidad Mayor, Santiago, Chile; Departamento de Física, Facultad de Ciencias, Universidad de Chile, Santiago, Chile

## Abstract

During the current COVID-19 pandemic it is imperative to give early warnings to reduce mortality. However, non-specialist such as authorities and the general population face several problems to understand the real thread of this pandemic, and under/overestimation of its risk are a commonplace in the press and social media. Here we define an index, which we call the COVID-19 Burden Index, that relates the capacities of the healthcare system of a given country to treat severe and critical cases. Its value is 0 if there is no extra strain in the healthcare system, and it reaches 1.0 when the collapse is imminent. As of 23 March 2020, we show that Chile, the USA, UK, among other countries, must reduce the rate of infections right now, otherwise, in less than 7 days they could be in a catastrophic situation such as Italy, Spain and Iran.

---

Is a nationwide quarantine, or other harder measures justified in Chile? The answer without deeper analysis is yes. But, the real tricky ingredient is timing. Scientific evidence has to be a key element in making this decision, and it has to be based on the available data. A reckless decision could do enormous harm to the national economy, which in Chile is already crippled by the social outburst of October 2019. However, acting too late is likely to imply a large number of causalities, or producing an even harder sociopolitical scenario. The long delay between the enforcement of measures and their repercussion in the infection rate makes taking the right decision even more difficult.

Our intention is to show, with just one -easy to grasp-parameter, how bad the situation facing the national healthcare system is. And, on that basis, infer if a nation-wide quarantine should be enforced as soon as possible. We avoid making forecasts about the number of infected, limiting ourselves to the available data. Our assumptions are minimal and play almost no role in our conclusions. We suggest a nationwide quarantine considering that our social life is very similar to that of Italy or Spain, who are experiencing a giant humanitarian crisis due to the collapse of their healthcare system. But, beyond this or another measure, we want to show that the contagion of the disease must be radically reduced right now, and this can be achieved by effective social distancing, only possible by lockdown.

## I. THE COVID-19 BURDEN INDEX

In an epidemic scenario, rather than the contagion percentage of the total population, one of the most critical elements is the strength of the healthcare system. In the COVID-19 pandemic, the bottleneck of the healthcare system is given by the number of beds *available* for intensive care (ICU). The number of ICU beds in Chile is not entirely certain, but it could be estimated to 1000 beds[1] (or *∼* 5.3 ICU per 100.000 inhabitants). Of those, we can expect a steady-state occupancy of 75%; this is the average of the usual hospital beds occupied in the OECD [2]. Due to the exponential growth of the infection, a lower value (*i*.*e*., a 50% or a 30% occupancy) does not make much difference. Therefore, for this study, we are going to consider the currently available capacity of the Chilean healthcare system for COVID-19 patients is *∼*250 ICU beds. As we will show later, a higher number of ICU beds can be assumed, for example, similar to the case of Belgium, but the scenario is still quite worrying.

The ratio of COVID-19 infected to those in need of intensive or critical care is about 15%, and a third of them require mechanical ventilation [3–6]. We will use 15% for our estimate. To compare the evolution in different countries we define the **COVID-19 burden** index as

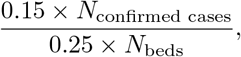

where *N*_confirmed cases_ the number of cumulative confirmed cases, and *N*_beds_ the number of ICU beds available in each country prior to 2020 [7]. A zero value means the basal occupancy of the healthcare system, *i*.*e*., the healthcare system in steady-state before the year 2020. When this index reaches 1.0 (the Burnout Limit) the healthcare system is at its local limit. A value larger than 1.0 reflects, approximately, how many extra ICU and medical professionals are needed relative to the regular healthcare system situation (75% occupancy). The important message is to keep the COVID-19 burden index below unity. For values above one doctors will surely find themselves in the difficult humanitarian dilemma of who to care for, or even worse, who to save.

Figure 1 shows the COVID-19 burden index for several countries using the data available [8]. The results are consistent with what is expected; the worst affected countries, *i*.*e*., Italy, Iran, and Spain score 4.6, 3.6, and 3.6 in the last day considered for this report. UK and Germany are about to reach their limit. Germany has maintained a controlled COVID-19 Burden index for 16 days; that is, it has maintained its health system without collapsing. But 16 days below the burnout limit may be insufficient, if they do not manage to reduce the rate, it spreads urgently. Although the disease is overcome on average in 2 weeks, critically ill patients take between 3 and 6 weeks [9], so Germany seems still far from overcoming the emergency. The UK case is very worrying because it could collapse very quickly if it does not reduce the rate of infections as soon as 2 or 3 days, or they could face the worst-case scenario. South Korea is on its limit, its healthcare system is fully occupied and strained, but it is copping with the demand. Finally, USA and Chile are steadily approaching the limits of their healthcare systems; in about one week, they might not be able to cope with the demand if they don’t reduce the rate of contagion right now. An under/over estimate of the parameters used could mean at most one or two days in reaching the burnout limit. Close to reaching this limit, the fatality of COVID-19 increases drastically, see Fig. 2.

**FIG 1.**
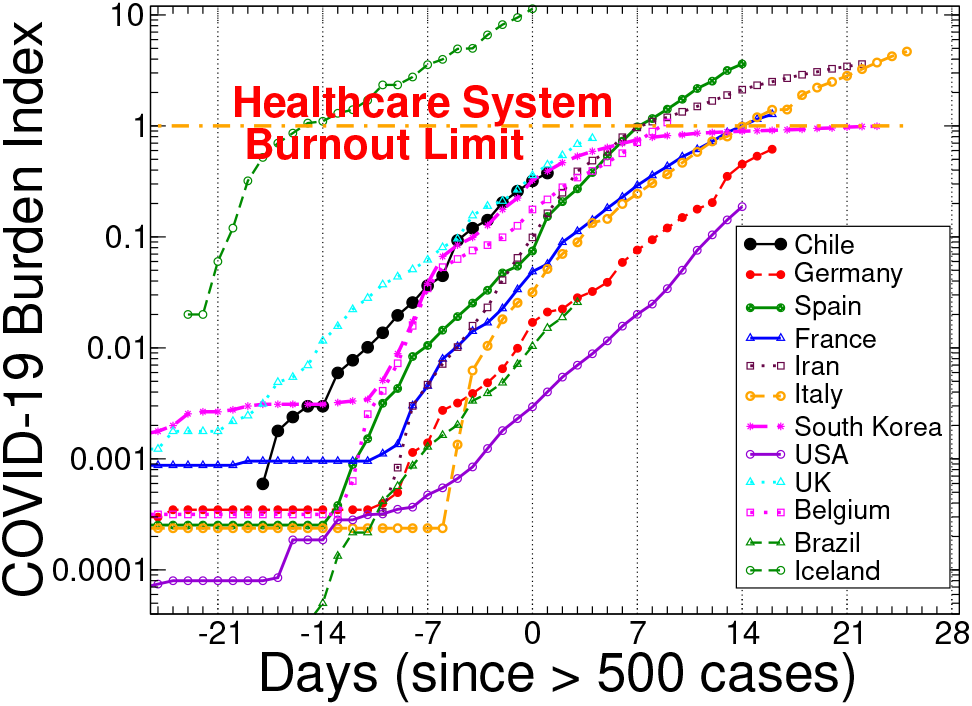
COVID-19 burden index for several countries as a function of time, after the first 500 positive cases are confirmed, a reasonable value for Chile. Notice that we present the index on a logarithmic scale.

**FIG 2.**
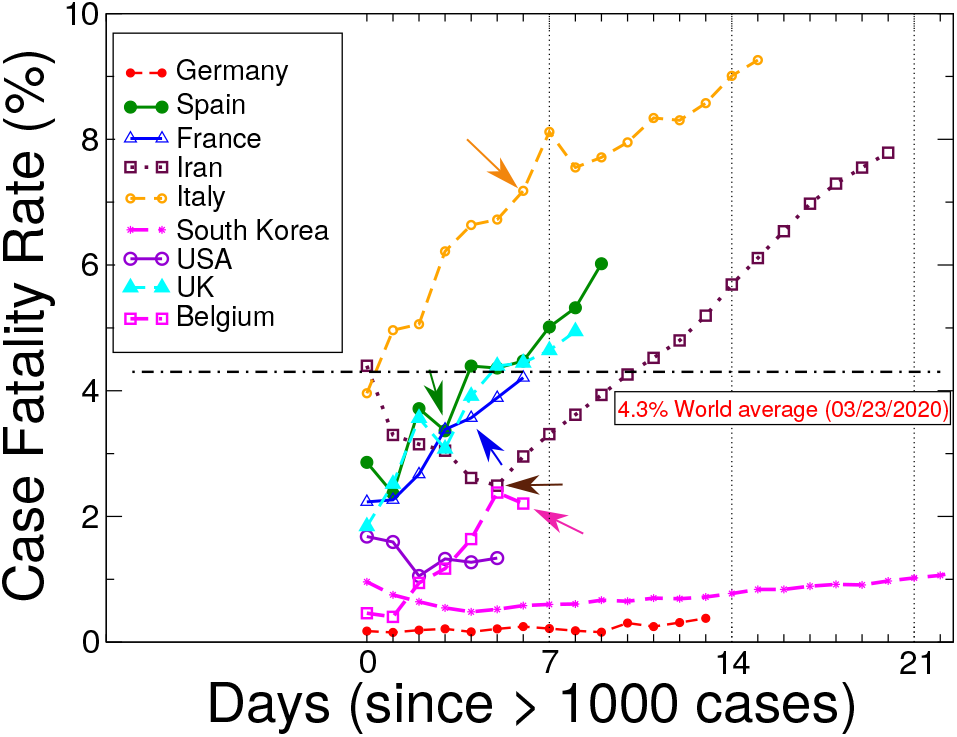
Fatality rate of COVID-19 by country. Only the data for countries with more than 1000 infected cases is provided. The black line is the world average by March 23, 2020. The small arrow indicates when the Burnout Limit was reached (COVID-19 Burden Index of 1).

Looking to the slope of the COVID-19 burden index in a logarithmic scale from countries such as Iceland, Belgium, Chile, France, Germany, and USA, all of them have similar behavior. At *early* stages, the contagion depends much on details, such as population, demographics and/or geography. Then, at *intermediate* stages -when around the 500 detected cases is reached-the slope of the COVID-19 burden index is nearly the same. This applies to countries so different as Iceland, Chile, USA and South Korea. It seems that once the exponential growth of the epidemic is unleashed, specific details for each country become irrelevant. Also, this slope does not correlate with the contagion rate found during the early propagation stages. Afterward, at *late* stages, the slope decreases. This is the case of South Korea, Italy and Iran. This third stage, we infer, is related to contagion control reducing the contagion rate. But, if this is accomplished too late as in Iran or Italy, the consequences are disastrous, as is being experienced. Our recommendation, for any country that is still in time, is to stay under the burnout point (1.0) as South Korea achieved. As we will show below, this can make a difference in reducing the number of deaths, when compared to the Italian crisis by about 10 times, using the preliminary data from South Korea, or even up to about 20 times as preliminary data from Germany. Finally, the marked difference, between 3 clear stages (or more in the case of South Korea) makes a prediction of the spread of COVID-19 inaccurate using standard methods, such as a regression of early contagious data (see Fig. 3).

**FIG 3.**
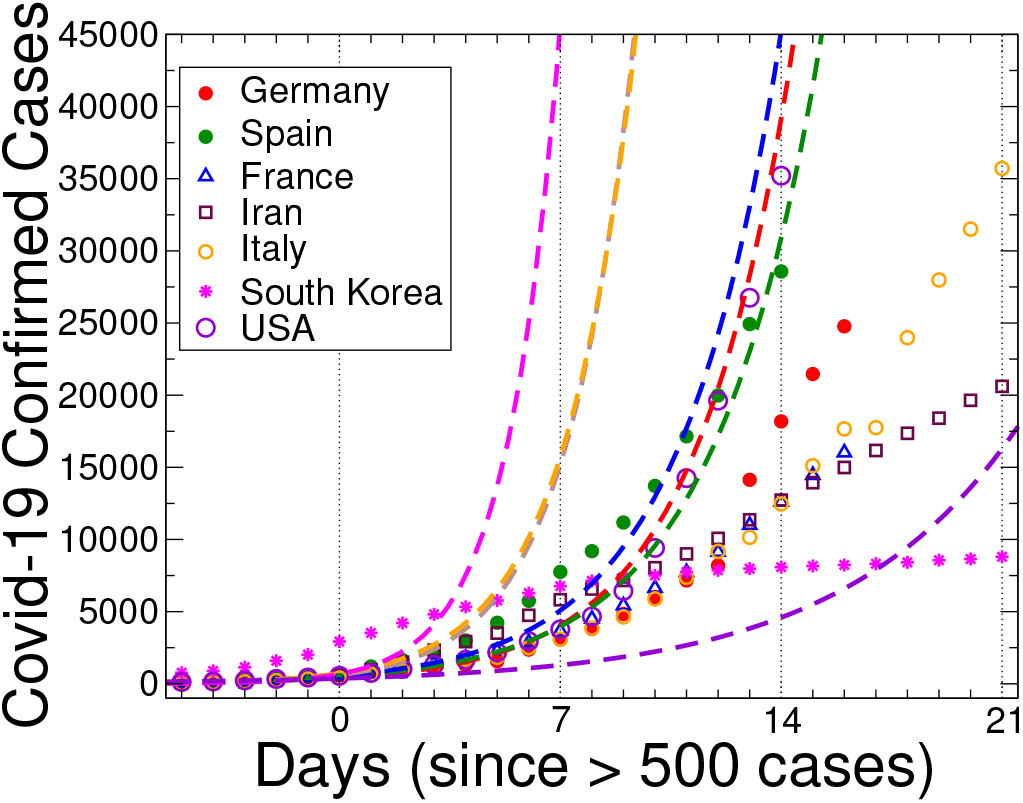
Is it possible to predict the contagion trend with data of about 500 cases? An exponential trend (dashed lines) was adjusted for each country real data using values over 50 and just over 500 cases. As a comparison, the actual data for the full period is plotted as symbols. Despite the common belief of decision-makers and the general population, it is NOT possible to give a truthful prediction, for better or for worse scenarios, during the early stages of COVID-19 spread based exclusively on the number of confirmed cases.

The most significant danger of this pandemic is not how contagious it is, or the mortality rate, which are important. The main danger is the collapse of the healthcare system. The case fatality rate (CFR), defined as the percentage of death from confirmed COVID-19 cases, changes day to day, and cannot be determined with certainty, and our data are no reference. However, on the basis of the data available on March 23, it is around 4,3%. This is in sharp contrast with South Korea, with over 8,000 cases and about 1% CFR. However, Spain and Italy, with their collapsed healthcare systems, report around 6 and 9%, respectively. Germany has not overcome the emergency and must take extreme measures. In any case, having today a similar number of confirmed infections than those of Spain, and after a similar time since they both exceeded 500 cases, it has a CFR of less than 0.5%, which seems encouraging.

Mainly, our intention is to sound an alarm. It is vital that the authorities of every affected country, on the basis of ICU available beds and the contagion rate, make their most to keep the COVID-19 Burden Index below 1, and take the severe and difficult decisions that this objective requires. In this analysis, we have tried to avoid any bias, sticking to the available data [8]. We believe that the COVID-19 burden index is a simple and solid tool to warn the authorities and decision-makers about the dangers of COVID-19. If the COVID-19 burden index reaches values close to 1.0 the collapse of the healthcare system is imminent, and harsh measures are in need to avoid a drastic increase in causalities.

Finally, while our main concern is Chile, the US case is very worrisome. In fact, with more than 100,000 ICU beds, twice as much as China, they are not shielding the population from exponential COVID-19 growth. The US has 17 times more population than Chile, and 100 times more ICU beds, but there is no guarantee of avoiding collapse (see Fig. 1). The only way to avoid collapse is by reducing the contagion rate as early as possible. By March 23, the slope of the COVID-19 burden index of USA is similar to South Korea in Day 2 (see Fig. 1). South Korea shortly afterward decreased the slope of the curve, avoiding the burnout limit. The same could happen in USA, but it will depend on the measures taken in the previous weeks.

The authors of this brief communication are used to analyze data and perform simulations in physics, trying to describe Nature. Regrettably, in this urgent communication, we rely only on real data and try to find a simple interpretation of a global humanitarian crisis to help on its solution. The scientific research using computational simulations and experiments are essential to stop this pandemic. Nevertheless, for this communication, the computational simulations were not necessary at all; the evidence from several countries is overwhelming. Unless the exponential growth is hindered in the following days, Chile, USA, and other countries in a similar situation could experience a similar collapse of their healthcare systems. Sadly, to stop the number of critical cases during the next week, the measures should have been taken about one week before writing this communication.

## Data Availability

No extra data available

## ACKNOWLEDGMENTS

The authors are supported by ANID Chile through Fondecyt grants Nos. 1191351, 1191353, and 11180557, the Center for the Development of Nanoscience and and Nanotechnology CEDENNA AFB180001, and Conicyt PIA/Anillo ACT192023.

